# MR Image Harmonization with Transformer

**DOI:** 10.1101/2023.08.16.23294184

**Authors:** Dong Han, Rui Yu, Shipeng Li, Jing Wang, Yuzun Yang, Zhixun Zhao, Yiming Wei, Shan Cong

**Affiliations:** College of Intelligent Systems Science and Engineering, Harbin Engineering University, Harbin 150001, China; Qingdao Innovation and Development Center, Harbin Engineering University, Qingdao 266520, China

**Keywords:** MRI, harmonization, Transformer, self-attention, hippocampus segmentation, ADNI

## Abstract

Many clinical applications require medical image harmonization to combine and normalize images from different scanners or protocols. This paper introduces a Transformer-based MR image harmonization method. Our proposed method leverages the self-attention mechanism of the Transformer to learn the complex relationships between image patches and effectively transfer the imaging characteristics from a source image domain to a target image domain. We evaluate our approach to state-of-the-art methods using a publicly available dataset of brain MRI scans and show that it provides superior quantitative metrics and visual quality. Furthermore, we demonstrate that the proposed approach is highly resistant to fluctuations in image modality, resolution, and noise. Overall, the experiment results indicate that our approach is a promising method for medical image harmonization that can improve the accuracy and reliability of automated analysis and diagnosis in clinical settings.

## I. Introduction

Magnetic resonance imaging (MRI) plays an essential part in aiding healthcare professionals with diagnostics and intervention by assisting in the identification of various diseases that affect different bodily systems, such as tumors, inflammation, trauma, degenerative diseases, and congenital diseases [1]. However, MR images captured by various MRI scanners may exhibit significant variations in intensity and other image characteristics. Moreover, the continued advancements in imaging protocols, biomarkers, and clinical measurements can lead to updates in the characteristics of the MRI data, thereby resulting in the production of different MR images for the same patient when scanned at different medical sites. Such variability, known as confounding information, poses a significant challenge for radiologists to evaluate images and for researchers to develop deep learning algorithms to build models from multi-site MRI image data [2].

One effective approach for mitigating the confounding information introduced by scanners and other variables is through the use of an MRI harmonization algorithm [3]. While traditional style transfer methods utilizing neural networks (CNNs) have been shown to represent the image style well, they have problems when attempting to separate image content from the style. To overcome this issue, several harmonization methods have introduced self-attention mechanisms to figure out the tasks.

However, CNN, on the other hand, has difficulty in capturing the relationships between long-distance features when it does not have sufficient layers. This is because of the restrictions in the receptive field that the convolution process imposes although deeper networks can be employed to overcome this limitation, this may lead to a reduction in details and features. Deeper networks can be utilized to circumvent this limitation; however, doing so may result in a reduction in details and features. In the process of convolutional feature extraction, losing features might have a negative effect on the content structure of the output. For example, on the edge of biological tissue, the transfer of information may become too rough. Moreover, these methods are unable to restore the original subtle features in MR images, thereby creating difficulties in downstream tasks such as segmentation and classification [4].

The latest development in artificial intelligence and machine learning has made it possible to use self-attention mechanisms in Transformer-based approaches [5], which can effectively learn the relationships between different layers of data [6] [3]. This contributes to a holistic understanding of global information [7], providing advantages in capturing the complex structures of visual data. However, due to the spatial and temporal coherence structure of visual data, a new network design is required to overcome the drawbacks of CNN-based methods. Transformer-based models have demonstrated success in various applications [8], such as image reorganization, object detection, and image generation [9]. This approach can facilitate the analysis of datasets consisting of images from diverse institutions and improve the generalizability of artificial intelligence models. Additionally, models trained on one dataset would be able to perform inference on datasets generated by scanners from different manufacturers, further enhancing the practical utility of this approach.

In this study, aiming at the content leak issue of CNN-based methods during the harmonization process, we present a new approach with Transformer architecture to address this problem. Unlike previous Transformer architectures, we introduce two transformer encoders to extract both style and content information from MR images, and two decoders to generate harmonized image patches. Additionally, changes in image resolution can significantly impact position encoding. Therefore, to capture semantic information in images of varying scales, we utilize a content-aware positional encoding scheme (CPE) method [10] that can effectively accommodate images of different sizes.

The primary objectives of our study are:

- To harmonize MR images and eliminate the confounding information introduced by various scanners and imaging protocols.
- To develop a novel Transformer-based model capable of adapting the imaging characteristics while restoring fine-grained details.
- To demonstrate the improved generalizability of machine learning algorithms in medical data through experiments.

## II. METHODOLOGY

Drawing inspiration from the success of style transfer in natural images, we adopt the method proposed by [10] and apply it to the MR imaging harmonization. This study proposes a content-aware positional encoding scheme method for accurately extracting positional information in MR images and further introduce a novel Transformer-based model to effectively harmonize these images. Fig. 1 shows the structure of the model.

**Fig. 1.**
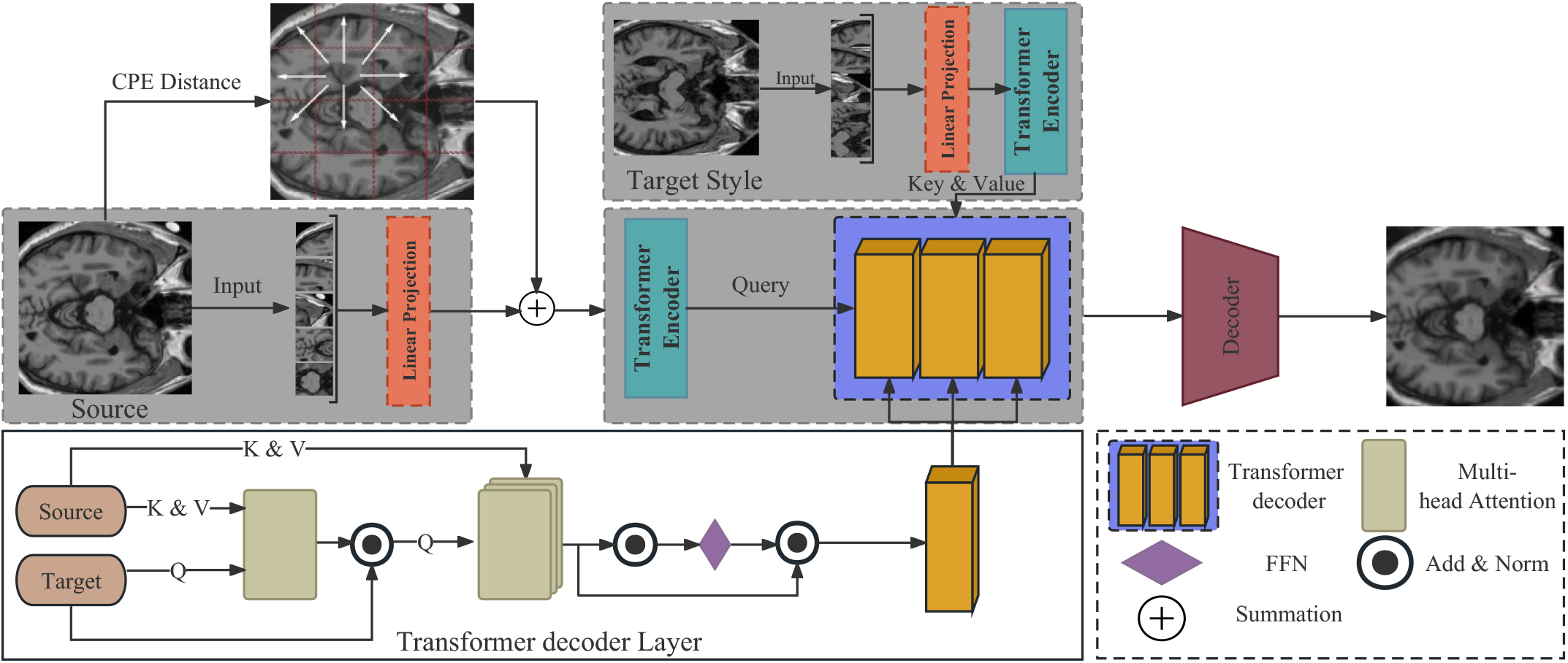
Our proposed harmonization framework is shown schematically above. Patches split from the input source pictures are fed into a linear projection layer to generate patch sequences. The target sequences are then processed by a style transformer encoder, while the source sequences, augmented with the content-aware positional encoding (CPE) distance, are fed into a transformer encoder. A decoder is utilized to generate the harmonized content from the input source sequences in the style of target sequences. An upsampling decoder is then used to reconstruct the unified picture at a higher resolution, significantly improving the output’s visual quality.

### A. Transformer Positional Encoding

Given the MR images are gray images, the channel *d* is fixed to 1, so the source and style image are respectively defined as *I*_*s*_, *I*_*t*_ ∈ ℝ^*H × W*^ then we split images into patches and map the patches into sequential feature embedding *ε*^*L × C*^, 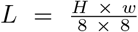, *C* represents the length of *ε*.

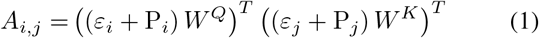

Positional encoding (PE) is used to extract structural information in the data. We derived the attention score calculation formula from [6]:

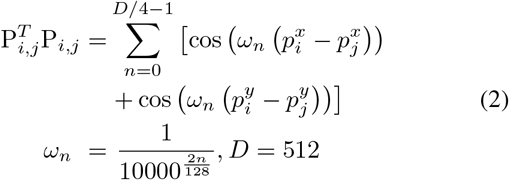

Where *A*_*ij*_ is the combined score for P_*i*_ and the P_*j*_ P_(*i,j*)_ is the positional encoding of P_*i*_ and the P_*j*_, *W* ^*Q*^ is parameter for query and *W* ^*K*^ is parameter for key. The encoding property of sinusoidal which makes sure distance between patches solely relies on their spatial distance [11]. However, the traditional positional encoding structure established for sentences is organized based on logic, image patches are arranged in content, and for more similar content patches, their distance should be less and they are supposed to have similar harmonization results. Meanwhile, for images of different scales, the relative distance between batches should not be affected.

To address this problem, we introduce content-aware positional encoding (CPE) to take the semantics of image content into account [10]. In detail, Given the input image *C* ∈ *ℝ*^*H × W*^, we use 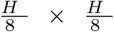 positional encoders to extract the content of the visual data. This approach ensures that the image resolution does not impact the spatial distance between patches, thereby enabling consistent and accurate patch-based processing of MR images.

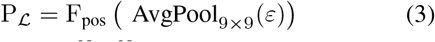

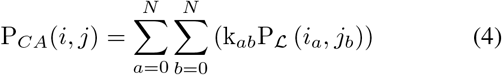

Where k_*ab*_ is the weight of interpolation patch and N is the number of nearby patches, P_*CA*_(*i, j*) is the CPE score for the P_*i*_ and the P_*i*_ at the position of (*i, j*), *AvgPool*_9 *×* 9_ is used to average pooling the (9 *×* 9) patch, F_*pos*_ is used to learn the content of the (1 × 1) patch. P_*ℒ*_ is used to learn the PE following the *ε*.

### B. Style Transformer

#### a) Transformer encoder

In contrast to previous networks based on CNN, we utilize two transformer encoders to separately encode content and style features, each encoder contains a multi-head self-attention module (MSA) and a feed-forward network (FFN). The incorporation of attention mechanisms enables us to capture global features, rather than just local information [6]. Specifically, convolutional operations derive information based on contextual content, while the attention mechanism splits data into queries (*Q* = *Z*_*C*_*W*_*q*_), keys (*K* = *Z*_*C*_*W*_*k*_), and values (*V* = *Z*_*C*_*W*_*v*_). By calculating the dot product between *Q* and *K*, attention coefficients are derived, which are then used to weigh the tokens. This approach effectively allows the model to focus on relevant features, while suppressing irrelevant or noisy information.

We encode the input content sequence *Z*_*C*_ = {*ε*_*c*1_ + P_*C*A1_, *ε*_*c*2_ + P_*C*A2_, …, *ε*_*cl*_ + P_*C*AL_ } and calculate the multi-head attention *F*_*MSA*_(*Q, K, V*) using the strategy in [12].

#### b) Transformer decoder

As displayed in Fig. 1, we apply the source image to calculate the *Q* and apply the target image to calculate the *K* and *V* . Now we need to regress the source sequence *Y*_*s*_ in the form of target sequence *Y*_*t*_.

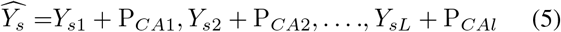

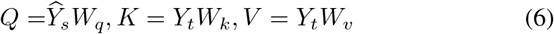

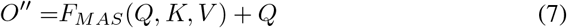

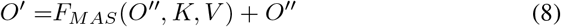

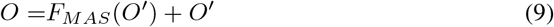

The output sequence *O* is calculated by the equation above. Where Y_*s*_ is the encoded source sequence. And behind each block, a normalization and addition layer is applied.

#### c) CNN decoder

we refine *O* using one-layer *CNN* decoder, which involves the following steps:

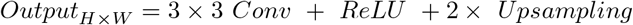

### C. Optimization function

Applying our Transformer-based model, the inference sequence should map both the source content and target style. In order to accomplish this, we design two perceptual loss functions: one is defined as the error in measuring content quality between the inference image and the source image, and the other is to evaluate the style quality of the inference image in comparison with the target image. To design the two loss functions, we utilize a VGG19 model that has been previously trained. Then we applied the Gram matrices method to calculate feature maps for the inference and target pictures and define style loss with it, while we apply the mean squared error (MSE) to calculate the error between the feature maps of the inference and the source image, and it is used to define the content loss. The use of these loss functions allows our model to effectively capture both the content and style of the input MRI images, thereby producing high-quality harmonized outputs [4], the content and style loss ℒ_*c*_, ℒ_*s*_ is defined as follows:

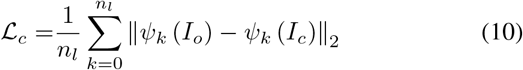

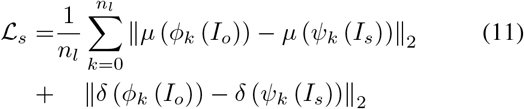

where *ψ*(*·*) represents the features captured from each layer in the VGG19 model, *µ*(·) and *δ(·*) stand for the mean and variance of extracted features, which has *n*_*l*_ layers. Finally, we apply the following loss function to optimize the outcome:

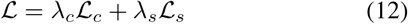

In our experiment, we set *λ*_*c*_ and *λ*_*s*_ to 15 and 9 to get the best performance [13].

## III. EXPERIMENTS AND ANALYSIS

In this part, we introduce a brief overview of MR image datasets used in our experiments, followed by a thorough description of the practical effects of the opposed approach and the experimental details.

### A. MRI Dataset

Accurate and sufficient data are critical to the success of artificial intelligence models. In order to meet this demand, we select MR image data from the Alzheimer’s Disease Neuroimaging Initiative (ADNI) dataset. The ADNI project, which was initially launched in 2004 as a public-private partnership, and led by the Principal Investigator Michael W. Weiner, MD. One primary aim of ADNI has been to examine whether serial imaging biomarkers extracted from MRI, positron emission tomography (PET), other biological markers, and clinical and neuropsychological assessment can be combined to measure the progression of mild cognitive impairment (MCI) and early AD. For up-to-date information, see www.adni-info.org. For our study, we choose the ADNI-1 project as our research dataset due to its completeness, accuracy, and extended duration. We select the EMCI research group as our dataset, consisting of 391 individuals. Confounding information in MRI scans is primarily introduced by the scanner used, as different manufacturers have their own production standards and scanners produce varying scan results. Therefore, we divide the data into four groups based on the MRI scanner manufacturer: SIMENS-1, SIMENS-2, Philips, and General Electric (GE). Of the 391 individuals, SIMENS-1 is scanned on SIMENS scanners with a 1.5 Tesla field strength, while the others are scanned on scanners with a 3.0 Tesla field strength. SIMENS-1 group comprises 90 samples, SIMENS-2 has 80, Philips has 104, and GE has 117 samples [14]

#### a) Data Labeling

In this study, we employ FreeSurfer, an open-source tool for analyzing neurogenesis data, to automatically label the MRI data. FreeSurfer offers a range of calculation methods to quantify the function, connectivity, and structure of the human brain. According to the research conducted by Zhong in 2010 [15], they compared five surface mapping of the cerebral cortex algorithms on 40 MR image scans. The results show that the segmentation result of FreeSurfer overlapped with the manual annotation by more than 80%. Thus, we use FreeSurfer to extract the left and right hippocampus area as our labels on 391 MRI Nifit data.

#### b) In the data labeling and pre-processing stage

we utilize the nibabel library in Python to convert the label results into a NumPy array with dimensions of 256 *×* 256 *×* 256. However, since the hippocampus is a relatively small region of the brain, there is a risk of inputting all-zero patches into the network during the training process, leading to low efficiency. To address this issue, we perform statistical analysis on the position of the hippocampus and found that it generally appears within the range of [70, 230], [80, 240], [120, 140] in 3 dimensions. Therefore, we crop the entire matrix to a size of 160 *×* 160 *×* 20, reducing the data scale to one-sixteenth of the original. We then extract 20 slices from each Nifit file by slicing the data along the z-axis. Finally, we save the segmentation matrix as NumPy data, which is an efficient format for model access.

### B. Evaluation Metrics

We evaluate and compare the opposed approach with three other proposed methods: AdaAttN [16], ArtFlow [17], and MAST [12]. AdaAttN performs attentive normalization on a per-point basis, ArtFlow consists of a pipeline based on neural flows and an unbiased feature transfer module, MAST is a typical CNN-based image harmonization approach, while our method employs transformer architecture to address the issue of losing fine details during the harmonization process. During the following two experiments, we quantify the improvement in picture quality and performance of hippocampal segmentation tasks based on the U-net method. For the content, we use the following metrics:

Feature Similarity (*FSIM*):

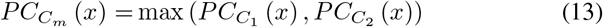

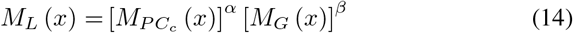

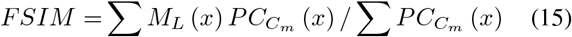

where, *C* ∈ ℝ^*H × W*^, *x* ∈ Ω.

Natural Image Quality Evaluator (*NIQE*):

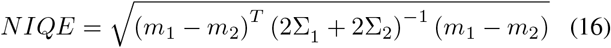

where *m*_1_, *m*_2_, Σ_1_, Σ_2_ denote the mean vector and covariance matrix of the natural and the distorted image MVG model.

Peak Signal-To-Noise Ratio (*PSNR*):

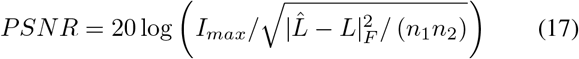

For the style, we calculate Universal Image Quality Index (*UIQI*) and Histogram Correlation Coefficient (*HCC*):

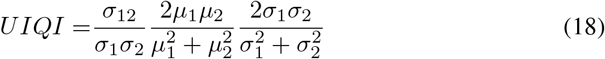

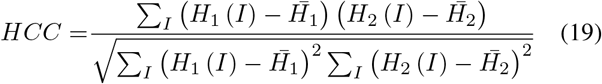

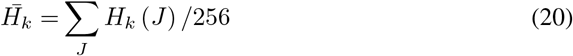

To evaluate the segmentation results, we employ three metrics: accuracy rate (*Acc*), intersection over union (*IoU*), and F1 score (*F* 1).

### C. Experiment 1: Style Transfer

#### a) Training details

The hardware used for this experiment consisted of an Intel(R) 6226R CPU @2.90GHz with 64 cores and an NVIDIA GeForce RTX 3090 with 24GB of memory. The operating system used is Cent OS 7 x86-64-7.1.1.

The overview of the experimental design is shown in Fig. 2, during the module training, the SIMENS-1 dataset (Field Strength = 1.5 Tesla) is used as the source dataset, and the SIMENS-2 dataset (Field Strength = 3.0 Tesla) is used as the target dataset. To enlarge the amount of training data, images in the source dataset are rotated by 90°, 180°, and 270°, resulting in a total of 5400 slices of source data. The algorithm is trained with 3780 SIMENS-1 slices, and the test set consists of the left 1080 slices, while 540 slices are utilized for validation. We apply Adam optimizer to make the training process more efficient, and the learning rate is set to 0.0002 [18]. A batch size of 4 is used, and the network is trained over 90,000 iterations, which lasts for about 10 hours. Fig. 3 shows the results of the testing. By computing the Universal Image Quality Index (UIQI), we compare the harmonization performance, the quantitative comparison of the different methods are shown in Tab. I and Tab. II.

**TABLE I.** Image quality evaluation on Philips set.

| Model | UIQI↑ | HCC↑ | FSIM↑ | PSNR↑ | NIQE↓ |
| --- | --- | --- | --- | --- | --- |
| Source | 0.029±0.0008 | 0.3±0.0033 | - | - | - |
| AdattN | 0.034±0.0011 | 0.5±0.0022 | 0.73±0.00051 | 59.9±8.5 | 18.87186±0.00016 |
| ArtFlow | 0.034±0.00107 | 0.36±0.0070 | 0.81±0.00026 | 64.7±23.1 | **18.8553±0.00023** |
| MAST | 0.033±0.00106 | 0.48±0.0037 | 0.77±0.0002 | 57.81±12 | 18.87726±0.00054 |
| Ours | **0.036±0.00116** | **0.58±0.0055** | **0.87±0.00044** | **70.45±28.06** | 18.86361±0.00078 |

**TABLE II.** Image quality evaluation on GE set.

| Model | UIQI↑ | HCC↑ | FSIM↑ | PSNR↑ | NIQE↓ |
| --- | --- | --- | --- | --- | --- |
| Source | 0.0284±0.00089 | 0.36±0.044 | - | - | - |
| AdattN | 0.0307±0.00098 | 0.53±0.051 | 0.75±0.00042 | 60.5±15.9 | 18.871±0.00024 |
| ArtFlow | 0.0287±0.00096 | 0.47±0.087 | 0.84±0.0002 | 62.3±27.0 | 18.865±0.00062 |
| MAST | **0.0317±0.00098** | 0.55±0.054 | 0.78±0.0001 | 58.9±15.2 | 18.876±0.00039 |
| Ours | 0.0300±0.001 | **0.65±0.080** | **0.89±0.00027** | **67.6±22.3** | **18.863±0.0001** |

**Fig. 2.**
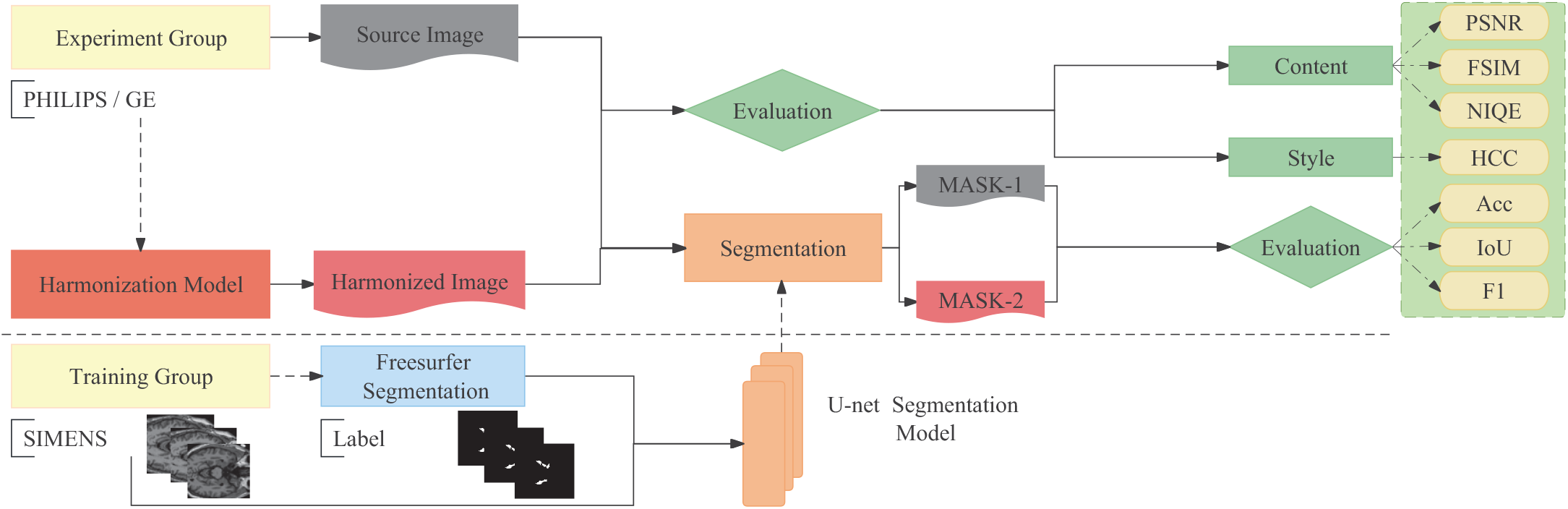
Overview of the experimental design. The U-net hippocampus segmentation model is trained using the SIMENS-2 dataset and corresponding labels. The hippocampus area in unharmonized and harmonized images is extracted by the segmentation model, and the resulting segmentation maps are evaluated to assess the performance of both the harmonization and segmentation tasks.

**Fig. 3.**
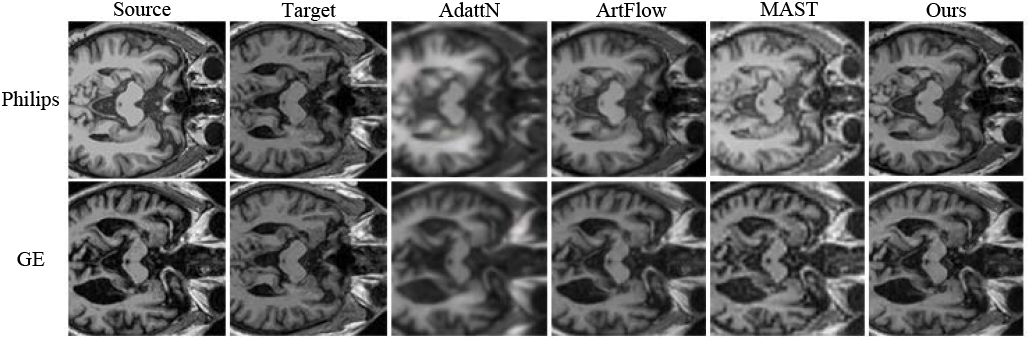
Harmonized MR images using different methods.

#### b) Quantitative evaluation

In this part, Philips and GE sets are employed as the source set and the SIMENS-2 set as the target set and fed the network to obtain the harmonized result. We analyze the structure and texture differences between the harmonized results and the input source images using FSIM, and we calculate PSNR and NIQE to evaluate the image quality. We calculate HCC to compare the correlation between the harmonized results and the input target image to evaluate the correlation of image characteristics. Tab. I and Tab. II illustrate the corresponding quantitative results. Overall, in most comparison metrics, our approach outperforms all other methods. Specifically, we achieve HCC values of 0.58 and 0.65 on the Philips and GE sets respectively, indicating successful alignment with the image characteristics of the target domain. In terms of preserving structure and texture information, the proposed method secures high FSIM scores, underlining its capacity to restore fine details and maintain tissue structures. Concerning image quality comparisons, our method excels in metrics like PSNR and NIQE, clearly demonstrating that image quality is enhanced during the harmonization process.

### D. Experiment 2: Hippocampus Segmentation

In the second experiment, we train a U-Net model [19] [20] to segment the hippocampus from other organisms, aimed at verifying the improvement of harmonization to the data. The model slices the raw data into patches of size [16,16] as input, with a stride length of [4,4]. With a learning rate set to 0.0002, and as the segmentation is based on the binary classification problem of gray scale [13], the input and output channels are both set to 1 [21].

#### a) Training details

In this experiment, we apply the same data augmentation technique as in experiment 1. We train the model with a training set of 3360 slices from the SIMENS-2 dataset. The test set consists of 960 slices, and the validation set has 480 slices. Next, we got the hippocampus segmentation results by feeding the harmonized experiment 1 result into the segmentation model, which is displayed Fig. 4.

**Fig. 4.**
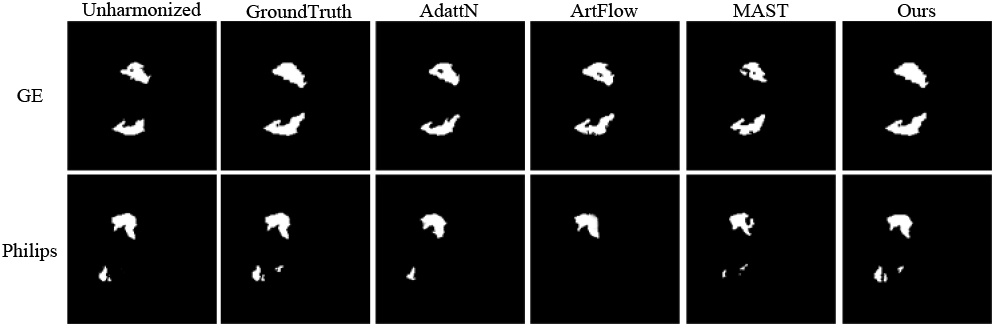
Comparison of hippocampus segmentation generated from our harmonized MR images and other harmonization methods.

#### b) Quantitative evaluation

We calculate the accuracy (Acc), Intersection over Union (IoU), and F1 score to evaluate the performance of our method on a hippocampus segmentation task. Fig. 5 and Fig. 6 show that our harmonization method, as demonstrated by evaluation metrics (Acc, IoU, and F1) on the Philips and GE datasets, enhances the performance of the existing segmentation algorithm compared to other results. Our method also exhibits fewer outliers, highlighting its robustness and stability. The conducted experiments affirm the efficacy of the proposed method in harmonizing MR images and enhancing the generalizability of machine learning models.

**Fig. 5.**
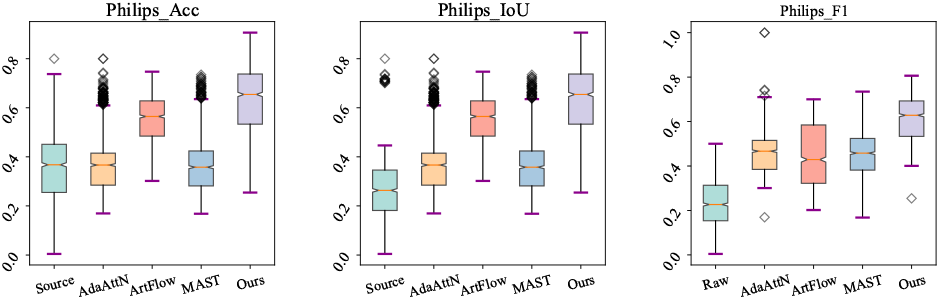
Segmentation evaluation of the harmonization results on Philips dataset.

**Fig. 6.**
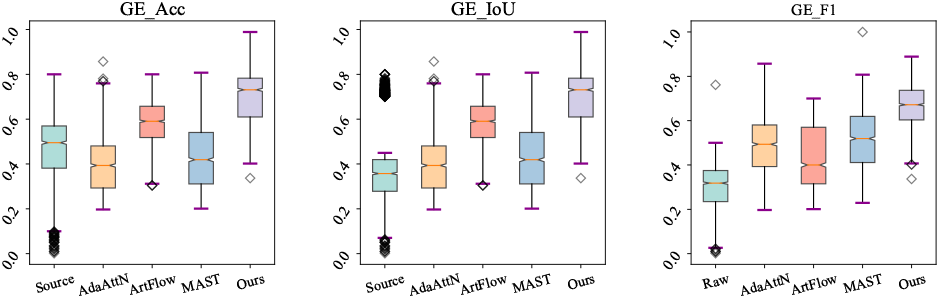
Segmentation evaluation of the harmonization results on GE dataset.

## IV. CONCLUSION AND FUTURE WORK

In this study, we propose a novel Transformer-architecture based MRI harmonization framework to accommodate MR images from diverse scanners and protocols. This framework treats image harmonization as a style transformation task where the imaging characteristics of source images are transferred to the target image domain via the decoder. Leveraging attention mechanisms, our model captures global properties beyond mere local information. Furthermore, we introduce a semantics-aware positional encoding method to overcome issues arising from varying image resolution. Our experiments demonstrate that our method aligns the characteristics of the source images and target images, preserving the original tissue structures and enhancing image quality. The experimental results from hippocampus segmentation further demonstrate that our harmonization method has promising potential to improve the generalization of machine learning models. Acknowledging the limitations of our study, we plan to perform further tests using diverse machine-learning algorithms and aim to extend our model to 3D volumes.

## Data Availability

All data produced in the present work are contained in the manuscript

## Funding Acknowledgments

This work is supported by NSFC 62103116 and Heilongjiang Provincial NSF LH2022F016.

## ADNI Data Acknowledgments

ADNI data collection and sharing for this project is funded by the Alzheimer’s Disease Metabolomics Consortium (National Institute on Aging R01AG046171, RF1AG051550, and 3U01AG024904-09S4).

## References

[1] M. Vockley, “Game-changing technologies: 10 promising innovations for healthcare,” Biomedical Instrumentation & Technology, vol. 51, no. 2, pp. 96–108, 2017.

[2] G. Modanwal, A. Vellal, M. Buda, and M. A. Mazurowski, “Mri image harmonization using cycle-consistent generative adversarial network,” Medical Imaging 2020: Computer-Aided Diagnosis, vol. 11314, pp. 259– 264, 2020.

[3] B. Neyshabur, S. Bhojanapalli, D. McAllester, and et al., “Exploring generalization in deep learning,” Advances in Neural Information Processing Systems (NeurIPS), 2017.

[4] X. Huang and S. Belongie, “Arbitrary style transfer in real-time with adaptive instance normalization,” IEEE International Conference on Computer Vision (ICCV), pp. 1501–1510, 2017.

[5] L. A. Gatys, A. S. Ecker, and M. Bethge, “Image style transfer using convolutional neural networks,” Proceedings of the IEEE conference on computer vision and pattern recognition, pp. 2414–2423, 2016.

[6] A. Vaswani, N. Shazeer, N. Parmar, J. Uszkoreit, L. Jones, A. N. Gomez, L. Kaiser, and I. Polosukhin, “Attention is all you need,” Advances in Neural Information Processing Systems, vol. 30, 2017.

[7] S. Paul and P.-Y. Chen, “Vision transformers are robust learners,” Proceedings of the AAAI Conference on Artificial Intelligence, vol. 36, no. 2, pp. 2071–2081, Jun. 2022.

[8] Z. Guo and D. Guo, “Image harmonization with transformer,” IEEE/CVF International Conference on Computer Vision (ICCV), pp. 870–14 879, 2021.

[9] X. Zhu, W. Su, L. Lu, B. Li, X. Wang, and J. Dai, “Deformable detr: Deformable transformers for end-to-end object detection,” International Conference on Learning Representations (ICLR), 2021.

[10] Y. Deng and W. Tang, “Stytr2: Image style transfer with transformers,” 2022 IEEE/CVF Conference on Computer Vision and Pattern Recognition (CVPR), pp. 11 326–11 336, 2022.

[11] Z. Yang, Z. Dai, Y. Yang, J. Carbonell, R. R. Salakhutdinov, and Q. V. Le, “Xlnet: Generalized autoregressive pretraining for language understanding,” Advances in Neural Information Processing Systems, vol. 32, 2019.

[12] Y. Deng, F. Tang et al., “Arbitrary style transfer via multi-adaptation network,” Proceedings of the 28th ACM International Conference on Multimedia, pp. 2719–2727, 2020.

[13] Y. Weng, T. Zhou, Y. Gong, Y. Xie, and C. Liang, “Nas-unet: Neural architecture search for medical image segmentation,” IEEE Access, vol. 7, pp. 44 247–44 257, 2019.

[14] R. Pomponio and Erus, “Harmonization of large mri datasets for the analysis of brain imaging patterns throughout the lifespan,” NeuroImage, vol. 208, p. 116450, 2020.

[15] J. Zhong, D. Phua, and A. Qiu, “Quantitative evaluation of lddmm, freesurfer, and caret for cortical surface mapping,” Neuroimage, vol. 52, no. 1, pp. 131–141, 2010.

[16] S. Liu, T. Lin et al., “Adaattn: Revisit attention mechanism in arbitrary neural style transfer,” 2021 IEEE/CVF International Conference on Computer Vision (ICCV), pp. 6629–6638, 2021.

[17] J. An, S. Huang, Y. Liu, Y. Wang, J. Guo, Y. Liu, Y. Yang, and J. Huang, “Artflow: Unbiased image style transfer via reversible neural flows,” 2021 IEEE/CVF Conference on Computer Vision and Pattern Recognition (CVPR), pp. 862–871, 2021.

[18] D. P. Kingma and J. Ba, “Adam: A method for stochastic optimization,” arXiv preprint arXiv:1412.6980, 2014.

[19] M. Bateriwala and P. Bourgeat, “Enforcing temporal consistency in deep learning segmentation of brain mr images,” arXiv preprint arXiv:1906.07160, 2019.

[20] S. R. Hashemi, S. S. Salehi, D. Erdogmus, S. Prabhu, and S. Warfield, “Asymmetric loss functions and deep densely connected networks for highly imbalanced medical image segmentation: Application to multiple sclerosis lesion detection,” IEEE Access, vol. 7, pp. 721–1735, 2019.

[21] T. Fan, G. Wang, Y. Li, and H. Wang, “Ma-net: A multi-scale attention network for liver and tumor segmentation,” IEEE Access, vol. 8, pp. 179 656–179 665, 2020.

